# Community-Tailored One Health Educational Intervention to Enhance Knowledge and Practices for Zoonotic Disease Prevention in Rural Thailand: a Protocol for a Prospective Cluster Randomised Controlled Trial in Chanthaburi, Thailand (Saan Suk trial)

**DOI:** 10.64898/2026.07.16.26358293

**Authors:** Marina Treskova, Cassia Rocha Pompeu, Pirrahyah Puntumetakul, Sasithorn Chaiphonngam, Kate Bärnighausen, Uliana Kachnova, Keeratiporn Jutaviriya, Monchai Phongsiri, Joacim Rocklöv, Till Bärnighausen, Patcharin Lapanun, Hans J. Overgaard

## Abstract

**Background:** Zoonotic infectious disease risk arises at human–animal–environment interfaces where pathogen spillover can occur. Rural communities living in biodiverse settings may experience frequent contact with wildlife and shared environments through livelihoods, food practices, and economic activities. Reducing spillover risk and strengthening pandemic prevention requires both structural and individual-level change. Community-based interventions that promote awareness, risk perception, self-efficacy, pro-environmental behaviour, and safe coexistence with wildlife may support prevention by shifting behavioural determinants of zoonotic disease risk. The Saan Suk intervention was co-developed with rural communities in Thailand using a Human-Centred Design approach and is grounded in the Health Belief Model and One Health principles. The intervention is intended to be feasible, acceptable, and deliverable through Thailand’s established Village Health Volunteer (VHV) system.

**Methods:** This protocol describes a parallel-arm, cluster-randomised controlled superiority trial that will be conducted during July - October 2026, in Chanthaburi Province, Thailand. 24 villages will be equally randomised to the Saan Suk intervention or the current practice (control). In intervention villages, trained VHVs will deliver, once a week over four weeks, a multimodal One Health educational intervention designed to improve knowledge of zoonotic spillover, promote protective behaviours, reduce risky wildlife-related contacts, and support respectful coexistence with wildlife. Trained outcome assessment teams will conduct structured interviews with 42 adult participants per village, yielding a total sample size of 1,008 participants. The sample size was calculated for the primary outcome, accounting for clustering, with 90% power to detect a medium effect size (6 points on the 0–100 knowledge scale) at a significance level of 0.05, accounting for a design effect with an ICC of 0.028. The primary outcome is knowledge of zoonotic spillover, transmission pathways, risk factors, protective and risky behaviours, and safe coexistence with wildlife. Secondary outcomes include attitudes, self-efficacy, preventive and risky behaviours, and reported contacts with major local reservoir hosts. A structured questionnaire was developed, expert-reviewed, and piloted for the outcome assessment. Outcomes will be analysed using mixed-effects regression models with random effects for village and adjustment for relevant pre-specified confounders. Primary analyses will follow the intention-to-treat principle.

**Discussion:** This trial will evaluate whether a co-designed, VHV-delivered One Health educational programme can improve knowledge of zoonotic disease prevention and behavioural determinants in rural communities living in close contact with wildlife and shared ecosystems. If effective and feasible, Saan Suk could inform integration into routine VHV training and community-based zoonotic disease and pandemic prevention strategies.

**Trial Registration:** The Saan Suk trial is registered with the German Clinical Trials Register (DRKS). Registration ID: <u>DRKS00038582</u>; date of registration: 11 May 2026.

**Metadata:** 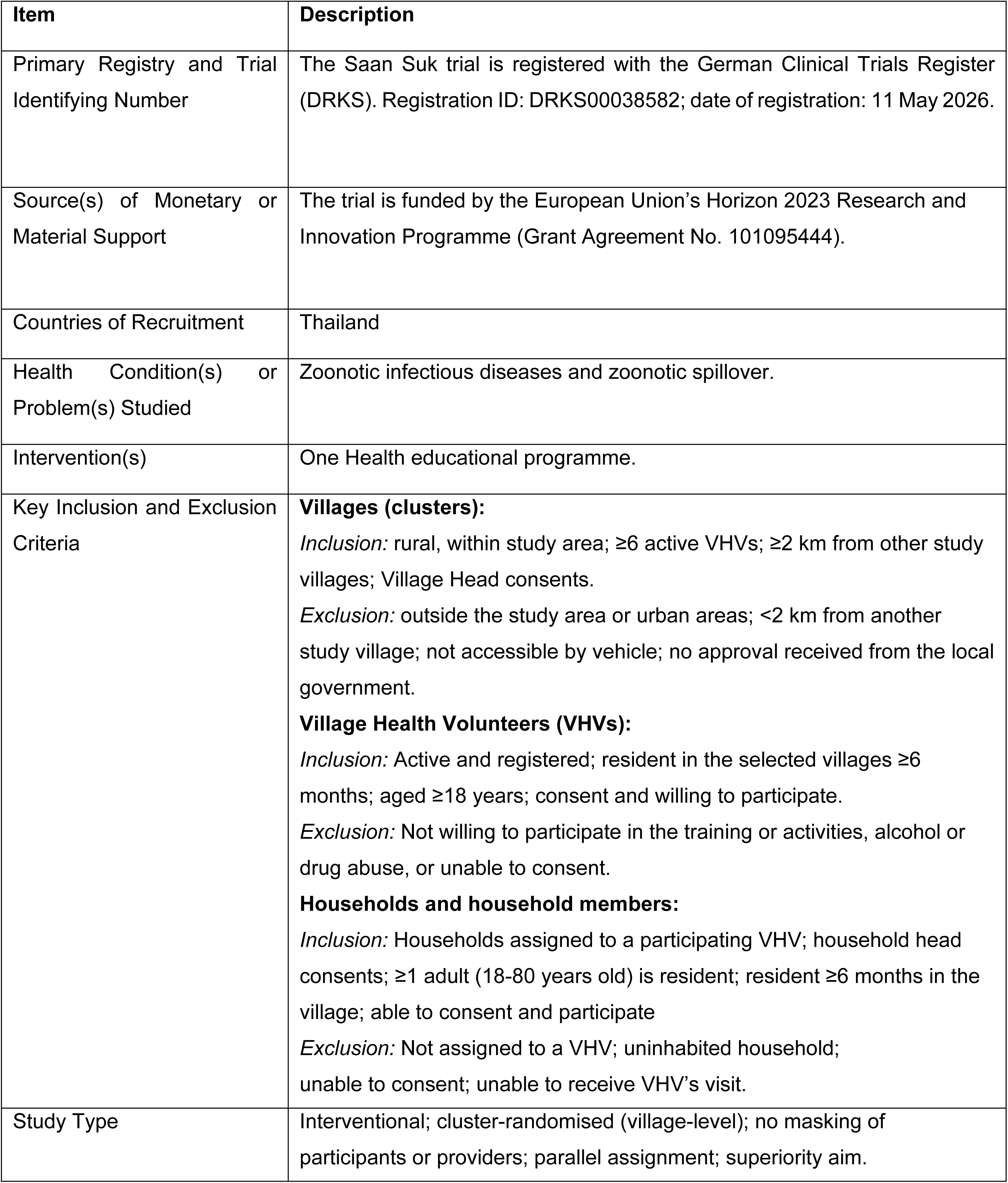

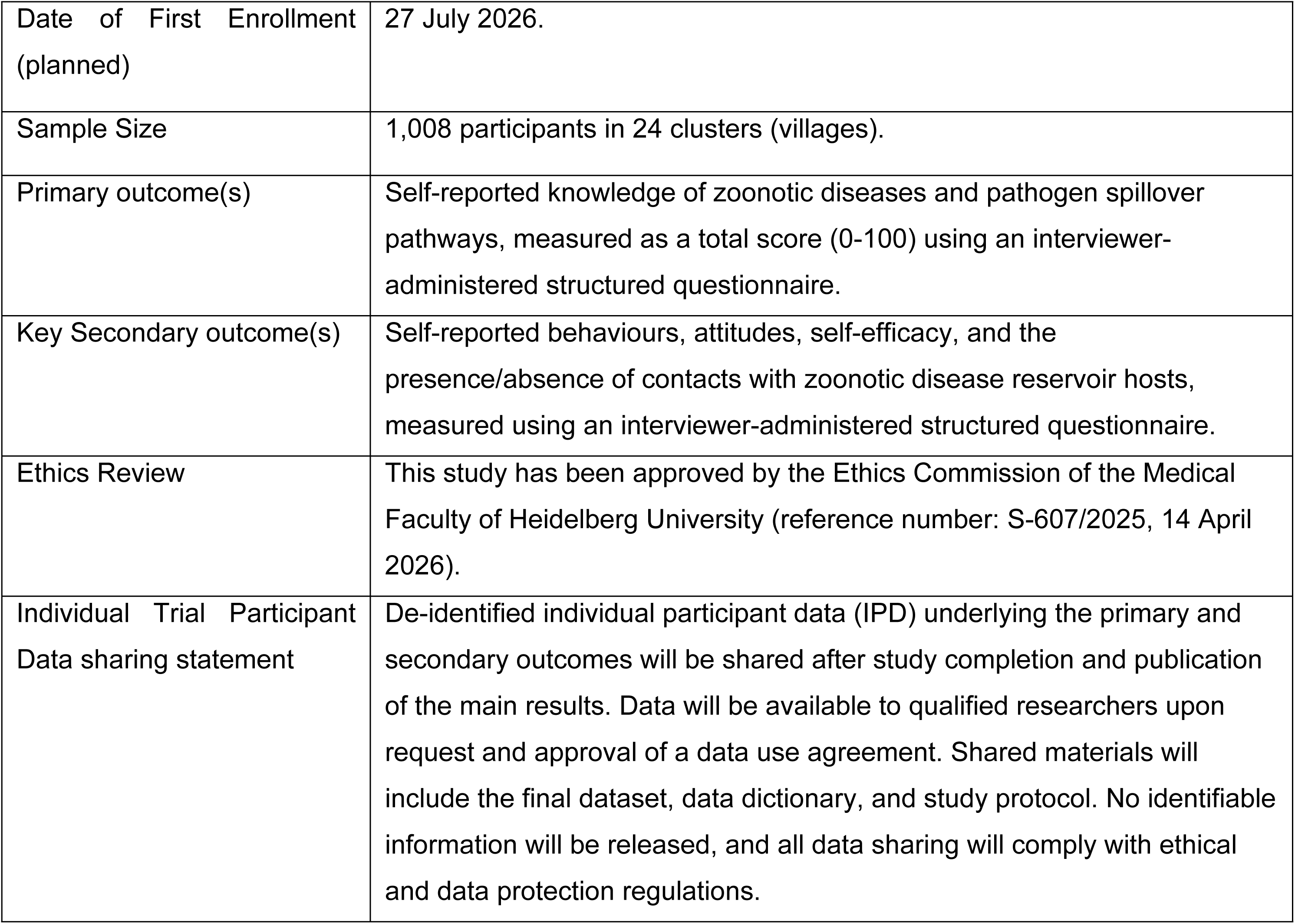

## Introduction

Zoonotic infectious diseases, transmissible from vertebrate animals to humans [1], can lead to large-scale outbreaks, as seen with COVID-19 (SARS-CoV-2) [2], the Middle East respiratory syndrome coronavirus (MERS-CoV) [3], and Influenza A (H1N1) [4], which generated severe impacts on public health and economy at a global scale [2]. Recent outbreaks of Andes Hantavirus linked to international cruise travel [5] and the ongoing Ebola Bundibugyo outbreak in the Democratic Republic of Congo and Uganda [6] further underscore the continued public health relevance of zoonotic infectious diseases and zoonotic pathogen spillover. Zoonotic spillover arises from a complex alignment of socio-ecological drivers at local and macro-ecological scales [7]. Human-driven changes, such as deforestation, habitat fragmentation, climate change, intensive farming, wildlife trade, and hunting [8,9], have contributed to increased human-animal interaction and exposure to pathogens, creating favourable conditions for zoonotic spillover [10]. In this context, One Health and EcoHealth approaches emphasise the interconnectedness of humans, animals, and the environment [11,12] and are essential for implementing effective zoonoses prevention, surveillance, and control measures [1]. Thailand has been recognised as a hotspot for biodiversity and human-wildlife interactions and is at risk of zoonotic disease emergence in Asia [13]. The country faces endemic zoonotic and vector-borne diseases (e.g., dengue fever, Chikungunya, Japanese encephalitis, simian malaria, and rabies) [14,15], and novel viruses identified in wildlife [16].

Community engagement interventions based on One Health approaches have effectively addressed public health challenges, especially in rural settings [17]. For instance, the “Lawa Project” [18,19] in Thailand combined school and community education, training of Village Health Volunteers (VHVs), ecosystem monitoring, and disease surveillance, significantly reducing opisthorchiasis in villages in northeast Thailand. VHVs were introduced into the Thai health system in the late 1970s to connect the formal health system with the community, becoming an internationally recognised public health model [20]. Managed by the Thai Ministry of Public Health (MOPH) under the Primary Healthcare Division, the programme encourages trusted community members living and working in the village to apply for VHV positions [21,22]. After acceptance, VHVs receive a brief training on primary health care, health promotion, disease prevention, and health education [20,22].

Thailand has over 1.1 million registered VHVs [23], the majority of whom are women (female-to-male ratio 4:1) aged 40 to 60 [24]. Each VHV is usually responsible for 7 to 15 households [24]. Their duties include informing, advising, and educating villagers on health issues and primary care, conducting routine screenings (e.g., malaria and blood sugar), collecting health data to inform national health programs, taking leading roles in community development and empowerment activities, and providing emotional and social support [20]. This multifaceted role positions VHVs as central actors in the healthcare system in Thai communities.

The effectiveness of community-based interventions relies heavily on individual behaviour, engagement, and adherence to preventive measures [25]. The Health Belief Model (HBM) provides a conceptual framework to investigate why an individual is motivated to take action to prevent or control health risks [26], and has been widely applied to studies such as adherence to COVID-19 precautionary measures in China [27], use of protective work equipment in Thailand [28], HIV prevention in Iran [29], and the zoonoses preventive practices by livestock farmers in Nepal [30].

Few studies, however, have integrated the HBM and One Health principles into community-based interventions. This integrated approach addresses both individual perceptions and behaviours and broader ecological and health factors, offering great potential to increase awareness and health education, ultimately reducing zoonotic pathogen spillover. Here, we describe a community-based One Health educational programme co-created with rural communities in Thailand, entitled Saan Suk (in Thai, “Weaving Health”), that addresses individual behaviour, health, and ecological drivers of zoonotic spillover risk and contributes to pandemic preparedness and prevention, and a protocol for a cluster randomised trial aiming to evaluate its effects. The programme was co-developed using a Human-Centred Design (HCD) approach [31], an iterative, participatory methodology that prioritises end-user needs, preferences, and contextual constraints, and is widely used in global health intervention development [32] and was piloted in the community. In this study, we aim to evaluate the effects of Saan Suk on targeted knowledge, behavioural, and attitudinal outcomes. This protocol describes the design and methods of a cluster-randomised controlled (cRCT) trial to assess these effects compared with current practice. If effective, the Saan Suk programme is intended to be suggested for integration into the existing national VHV training programme as part of routine training and community health promotion activities in Thailand.

## Materials and Methods

### Aims, study design, and settings

We will implement the Saan Suk cRCT in rural villages of Chanthaburi province, eastern Thailand. We hypothesise that community members in villages allocated to the Saan Suk intervention will show better knowledge and understanding of zoonotic spillover, transmission pathways, risk factors, perceived susceptibility and severity, and preventive behaviours compared with community members in villages continuing the current practice. Therefore, the primary objective of the Saan Suk cRCT is to evaluate whether the Saan Suk educational programme (intervention) improves community members’ knowledge and understanding of zoonotic spillover, transmission pathways, risk factors, perceived susceptibility and severity, and preventive behaviours, as informed by the Health Belief Model and socio-ecological drivers as informed by the One Health approach. The secondary objectives of the Saan Suk cRCT are to: 1) assess changes in attitudes, perceived susceptibility, and perceived severity of zoonotic disease risks, in line with the Health Belief Model; 2) assess whether the intervention changes self-reported preventive behaviours that reduce direct and indirect contact with wildlife, domestic animals, and potentially infectious animal tissues; and 3) evaluate whether hands-on training and skill-building delivered by VHVs enhances community members’ self-efficacy to translate knowledge into preventive practices.

The Saan Suk trial is a parallel-arm, cluster-randomised superiority trial evaluating the effectiveness of the Saan Suk intervention on the primary and secondary outcomes. Villages are the unit of randomisation and the Saan Suk intervention allocation. Villages will be allocated equally, i.e., using an 1:1 ratio, to the intervention or control arm using simple randomisation. The cluster design was selected to minimise intervention spillover between participants living in the same village and to reflect the planned mode of intervention delivery through village-level health structures.

The target population will be adult community members living in rural villages in Chanthaburi Province. Chanthaburi has approximately 536,000 inhabitants [33] spread across 10 administrative districts, with livelihoods primarily based on nature, and the local economy based on agriculture, fruit cultivation, and livestock raising. These social and ecological conditions make it a suitable setting for testing the effectiveness of the community-based One Health knowledge intervention aimed at preventing zoonotic diseases. The Saan Suk trial will be conducted in two districts of Chanthaburi: Pong Nam Ron District, including the sub-districts Thap Sai, Nong Takhong, Khlong Yai, and Thep Nimit, and Soi Dao District, including the sub-districts Thung Khanan, Saton, and Sai Khao. Both districts border Cambodia. Figure 1 presents the CONSORT flow diagram for the Saan Suk cRCT.

**Figure 1.**
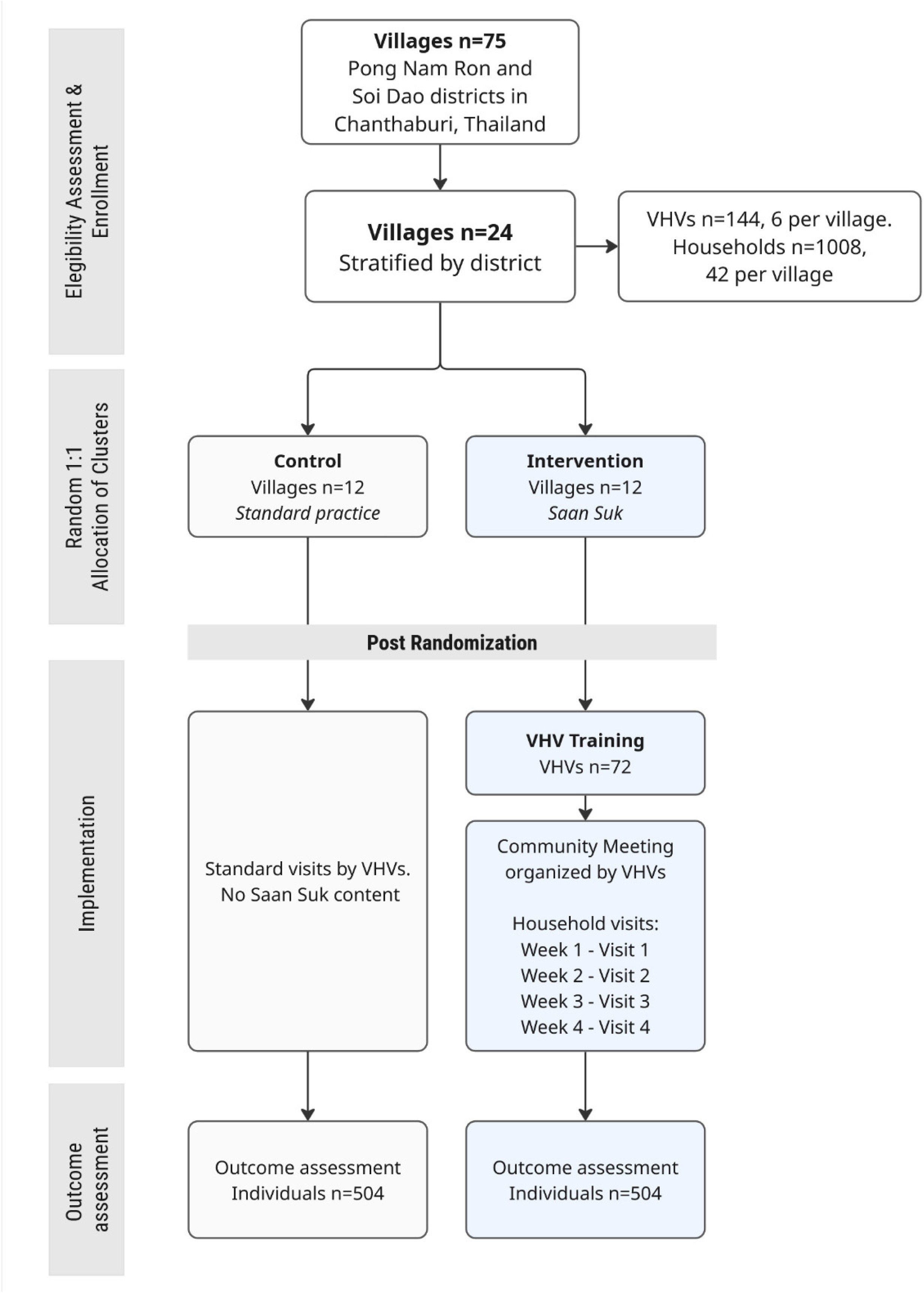
Trial flow diagram showing village randomisation, household recruitment, intervention delivery, and outcome assessment in the Saan Suk cluster-randomised controlled trial.

### Sample size calculation

The sample size for the Saan Suk cRCT was calculated based on the primary outcome, i.e., the knowledge score, scaled to 0-100. Parameters were derived from a cross-sectional survey conducted in Chanthaburi in November 2024 (n = 687, unpublished data), with a resulting mean knowledge score of 57.35 and standard deviation (SD) of 15.23. An intracluster correlation coefficient (ICC) of 0.028 was obtained by fitting a random-intercept linear mixed-effects model with village as the clustering variable. Assuming a two-sided significance level of α = 0.05 and 90% power, the trial is powered to detect differences in mean knowledge scores between the intervention and control arms at the post-intervention assessment. We parameterised the expected effect using Cohen’s *d* of 0.40 [34], corresponding to approximately six points on the 0–100 knowledge scale, based on the estimated SD from the cross-sectional survey. This effect size is considered a small-to-moderate improvement in educational intervention research, and it is viewed as a meaningful and achievable change in population knowledge over a short period.

Sample size adjustments accounted for clustering, a 15% allowance for both individual- and cluster-level attrition, and a 5% inflation to account for potential contamination. Based on these assumptions and operational feasibility, the final design includes 24 clusters (villages), with 12 clusters per arm, with an average cluster size of 42 participants (i.e., 6 VHVs per village, visiting 7 households each), resulting in a total sample size of 1008 individuals (504 per arm) (Figure 1).

### Participants

Villages (clusters) and VHVs delivering the intervention will be assessed for eligibility based on the following inclusion and exclusion criteria. Villages will be eligible if (1) they are classified as rural and located within the study area; (2) they have at least six active VHVs; (3) they are accessible by vehicle (a van); (4) they are located at least 2 km from other participating villages; (5) their Village Heads provide written informed consent; and (6) they are considered feasible for study implementation based on prior scoping visits and survey data. Villages will be excluded if they meet any of the following criteria: (1) located in urban areas or outside the study area; (2) not feasible for implementation due to security concerns (e.g., armed conflict, civil unrest) or lack of governmental approval; or (3) located less than 2 km from another participating village.

VHVs will deliver the Saan Suuk educational programme to participating households in their assigned villages. VHVs will be eligible if they meet all of the following criteria: (1) are currently active in their role and regularly serving at least seven households; (2) have been living in the selected village for at least six months prior to enrollment and intending to remain for the duration of the trial; (3) are registered in the national VHV registry; (4) participate in routine VHV training at the local hospital; (5) are aged ≥18 years; (6) are able to provide informed consent; and (7) are willing to participate in a 2-3 days training programme. VHVs will be excluded if they meet any of the following criteria: (1) are unable to provide informed consent; or (2) decline or withdraw consent at any stage of the study.

Households are eligible if they are permanent households in a participating village, are assigned to a VHV included in the trial procedures, have at least one adult household member aged 18–80 years, and the Household Head provides informed consent for household participation. Households are not eligible if they are uninhabited or not permanently occupied, are assigned to a VHV not included in the trial procedures, have no household member aged 18–80 years, or the Household Head does not provide consent.

Adult household members are eligible if they are 18–80 years old, legally able and mentally competent to provide informed consent, have lived in the village for more than 6 months before enrollment, can participate in the educational intervention and outcome assessments, and provide written informed consent. Adult household members are not eligible if they are younger than 18 years or older than 80 years, have lived in the village for less than 6 months, are unable to provide informed consent or meaningfully participate, do not speak Thai, are physically unable to attend study activities, are under the influence of alcohol or drugs during study procedures, or decline or withdraw consent.

### Recruitment

Before cluster enrollment begins, the trial coordination team will select the required number of clusters from the list of villages in each district, along with their geolocations. Villages will be randomly chosen from the eligible sampling pool, with the additional requirement that participating villages must be at least 2 km apart to reduce the risk of intervention spillover between the trial arms. To account for potential refusals or withdrawals, two additional villages per district will be preselected as replacements. Further, we will select the VHV Head and randomly five VHVs for each village using the government VHV registry, following the eligibility criteria. The VHV Head in each village will ensure study implementation. In each village, we will further pre-select two replacements in case selected VHVs decline to participate.

Before cluster and participant enrollment, we will conduct a scoping and engagement visit to secure the necessary institutional and local permissions, ensure alignment with national and provincial priorities, and facilitate stakeholder collaboration. We will recruit villages for the Saan Suk cRCT through existing district-level administrative structures. We will send official invitation letters from Khon Kaen University to the Chief of District supporting the study implementation and to the Village Heads of the selected villages, presenting the study objectives, procedures, risks, and benefits, as well as the list of selected VHVs for participation.

Further, we will invite the Village Heads and VHV Heads of the selected villages to an in-person meeting at the two district offices. During these meetings, the trial research team will introduce the objectives and procedures of the Saan Suk trial. Village Heads and VHV Heads who agreed, on behalf of their villages, to participate will be invited to sign informed consent forms.

Following the village and VHV Head enrollment, we will return to the villages to explain the meeting outcomes and the study implementation to the randomly selected VHVs from the registry and obtain their agreement to participate in the study using informed consent procedures. Participating VHVs will provide information on the households they are usually responsible for, including the number of people in each household, the number of eligible adults, their ages, and names. This data will serve as a sampling frame for the random selection of seven households to participate in the trial procedures. The number of households that a VHVs regularly visits can range from 7 to 15. The data manager in the coordination site will receive a digitised list of household IDs, including the names and ages of the adults. The data manager will then randomly select seven households for participation using a pre-developed function in *R* software that facilitates random selection of the needed number from the provided sequence, i.e., 7 from n = 1-15. The data manager will provide an additional two households for replacement in case the selected household does not agree to participate. VHVs will inform them about the study and obtain verbal consent to participate in the trial.

### Randomisation

Selected villages will be allocated to either the intervention or control arm using a 1:1 ratio through a transparent, non-computer-based ceremonial randomisation procedure. Randomisation will be conducted during official public meetings at the District Offices of Pong Nam Ron and Soi Dao; therefore, it will be restricted to the district level for operational feasibility. A total of 24 identical, opaque, sealed envelopes will be prepared in advance and used for allocation (intervention or control) by a member of the trial coordination team not involved in recruitment, enrollment, or intervention delivery. During the ceremony, Village Heads will draw sealed envelopes containing the pre-prepared allocation assignments. To maintain transparency and stakeholder engagement, Village Heads will draw one envelope each in the presence of the trial team, other Village Heads, VHV Heads, and District Chiefs. The allocation will be revealed after the VHVs in each arm have provided informed consent for participation in the trial. The allocation results will then be securely transmitted to the central trial data management team.

### Blinding criteria

Blinding will be implemented where feasible, considering the community-based nature of the intervention. Due to the design, it will not be possible to blind VHVs, participants, or the local trial team to allocation status. Outcome assessors will be blinded to the intervention allocation. Data analysts, including the statistician, principal investigator, and trial supervisors, will also remain blinded to allocation until completion of the primary analysis. The data manager will not be blinded, as access to allocation information is required for data linkage and management. Details of blinding implementation are provided in Table 1.

**Table 1.**
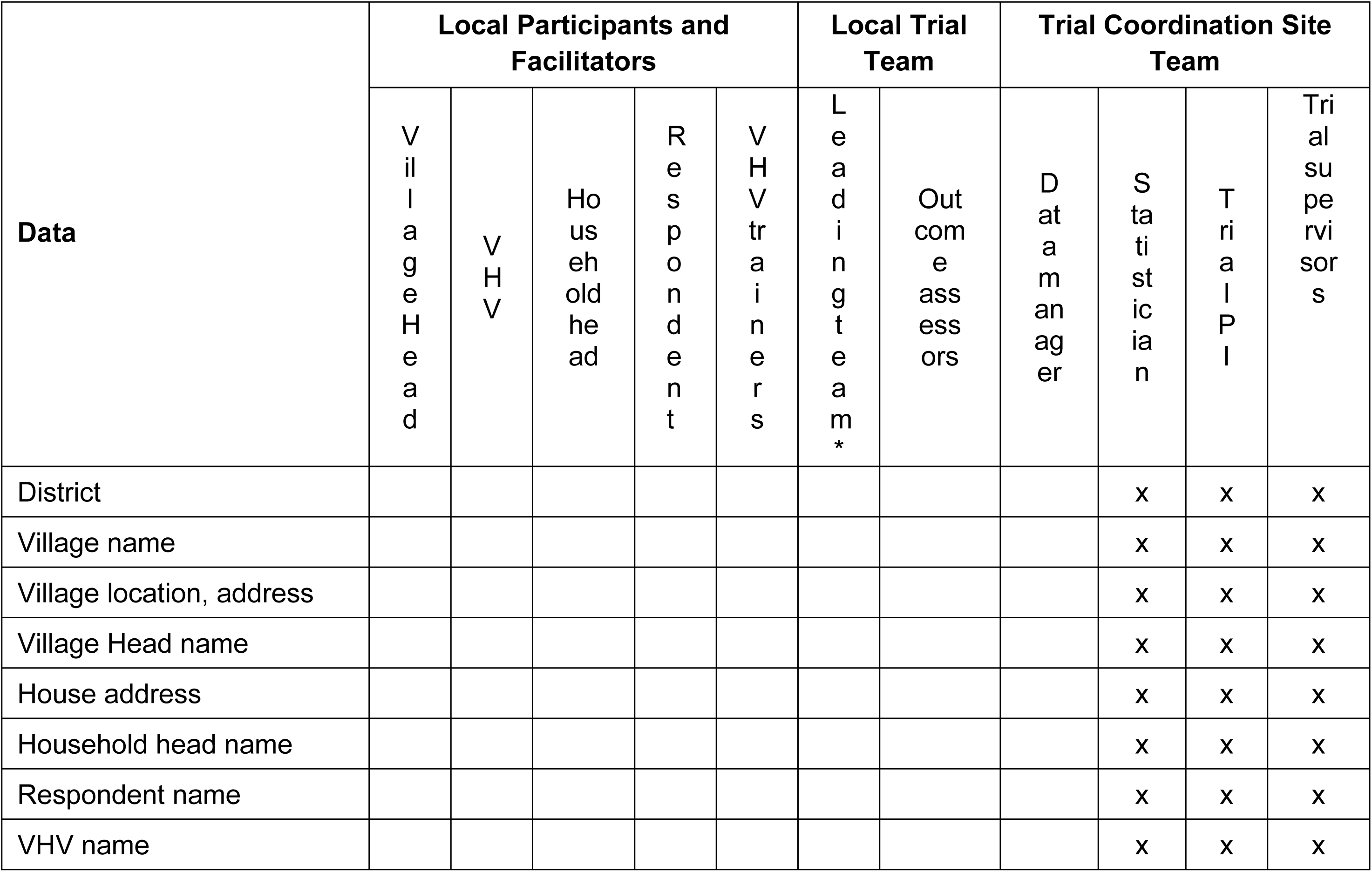
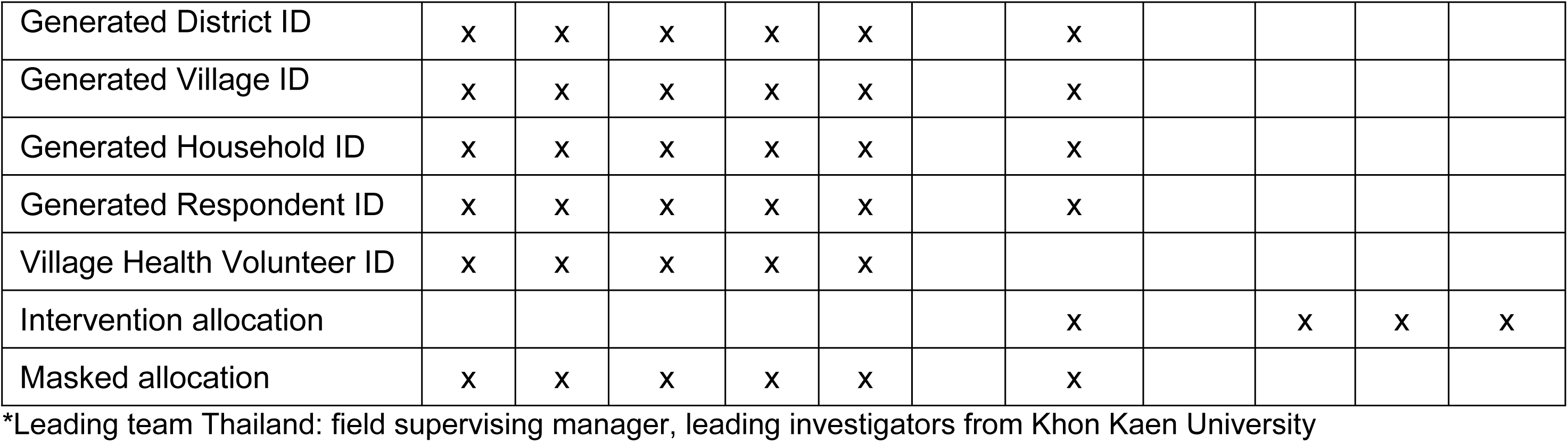
Implementation of blinding in the Saan Suk cRCT (ticked box indicates blinding to the information)

| Data | Local Participants and Facilitators |  |  |  |  | Local Trial Team |  | Trial Coordination Site Team |  |  |  |
| --- | --- | --- | --- | --- | --- | --- | --- | --- | --- | --- | --- |
|  | Village Head | VHV | Household head | Respondent | VHV trainees | Leading team* | Outcome assessors | Data manager | Statistician | Trial PI | Trial supervisors |
| District |  |  |  |  |  |  |  |  | x | x | x |
| Village name |  |  |  |  |  |  |  |  | x | x | x |
| Village location, address |  |  |  |  |  |  |  |  | x | x | x |
| Village Head name |  |  |  |  |  |  |  |  | x | x | x |
| House address |  |  |  |  |  |  |  |  | x | x | x |
| Household head name |  |  |  |  |  |  |  |  | x | x | x |
| Respondent name |  |  |  |  |  |  |  |  | x | x | x |
| VHV name |  |  |  |  |  |  |  |  | x | x | x |
| Generated District ID | x | x | x | x | x |  | x |  |  |  |  |
| Generated Village ID | x | x | x | x | x |  | x |  |  |  |  |
| Generated Household ID | x | x | x | x | x |  | x |  |  |  |  |
| Generated Respondent ID | x | x | x | x | x |  | x |  |  |  |  |
| Village Health Volunteer ID | x | x | x | x | x |  |  |  |  |  |  |
| Intervention allocation |  |  |  |  |  |  | x |  | x | x | x |
| Masked allocation | x | x | x | x | x |  | x |  |  |  |  |

\*Leading team Thailand: field supervising manager, leading investigators from Khon Kaen University

Routine unblinding will not occur during the trial. All outcome assessments and primary statistical analyses will be conducted with masked allocation, and the study database will remain blinded until database lock and completion of the prespecified primary analyses. Unblinding before database lock will be permitted only under exceptional circumstances in which knowledge of intervention allocation is required for ethical, regulatory, or legal reasons.

### Informed consent

We will obtain informed consent from four groups: Village Heads, Village Health Volunteers (VHVs, including VHV Heads), Household Heads, and adult Household Members selected for the outcome assessment. All individuals will receive a participant information sheet and will have the opportunity to ask questions before providing consent. Informed consent from Village Heads, VHV Heads, and VHVs will be obtained by the research team before the allocation of interventions. Village Heads will provide consent for their villages to participate in the study before randomisation. In the intervention arm, household members will provide informed consent before the intervention implementation. In the control arm, individuals participating in the outcome assessment will be asked to provide informed consent before the procedure.

### Intervention and comparator

The Saan Suk is a community-based One Health educational programme designed to be delivered by trained VHVs during routine household visits in rural Thailand. It was co-developed with community stakeholders in rural Thailand, including Village Heads, VHVs, and community members, using the HCD approach [31]. Community input and feedback guided the development of the intervention materials, delivery format, culturally appropriate content, and field feasibility considerations during the design and pilot phases. The Saan Suk programme is multimodal and includes community engagement, educational videos, leaflets, calendars, and practical items, including soap, biodegradable gloves, wet tissues, and cutting boards, used as learning tools to be delivered by VHVs to introduce and reinforce key information and support preventive behavioural practices. Different modes of the Saan Suk programme serve distinct but complementary functions integrated into each visit: 1) videos and leaflets establish risk identification and knowledge of transmission mechanisms; 2) practical demonstrations and handouts of protective items build self-efficacy and behavioral skills by providing tangible, actionable strategies; 3) calendars and take-home artefacts (such as cutting boards reinforcing food separation practices) serve as environmental cues and reminders that extend learning beyond the visit, supporting habit formation and maintenance of safer behaviours; and collectively, 4) these materials articulate the benefits of protective actions through concrete, contextualised examples relevant to household and agricultural settings (Figure 2).

**Figure 2.**
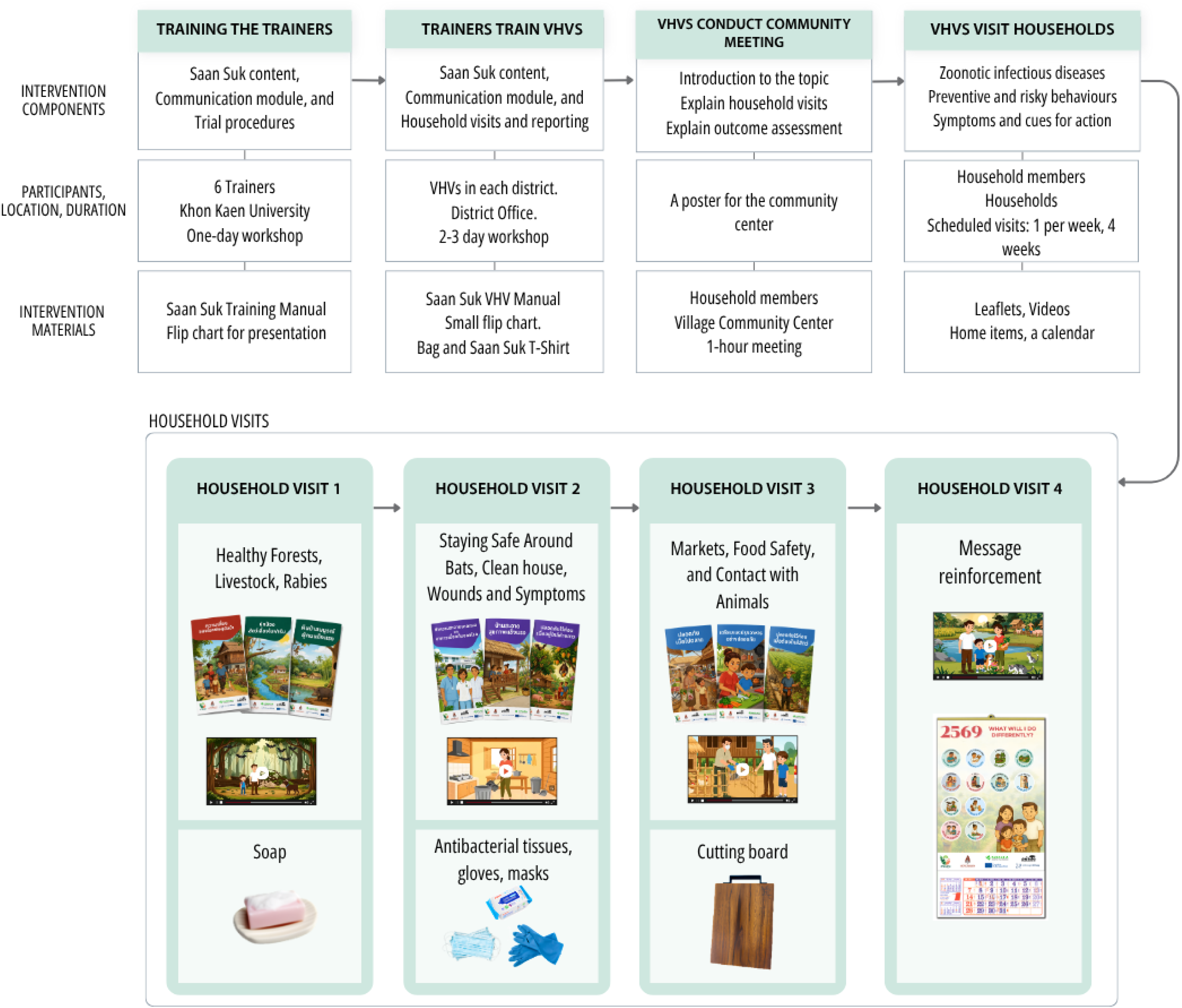
Schematic representation of the Saan Suk intervention components.

By combining informational, behavioural, and motivational elements across multiple modalities, the materials address the sequential pathway from risk awareness to behavioural intention and the adoption of sustainable practices. Adherence to the intervention will be supported through existing community health structures, structured supervision of VHVs, and endorsement by the Chief of District and provincial health authorities. VHVs maintain established relationships with households through routine visits, which is expected to facilitate engagement and continued participation in the intervention. VHV participation in the study will be limited to VHVs who are active in their role and willing to engage in training and all study procedures. Engagement of Village Heads will further support adherence at the community level, as their endorsement of the study and facilitation of the Saan Suk activities are expected to promote participation and sustained engagement among households. Intervention delivery will be monitored through activity logs maintained by VHVs, including records of household visits, dates, and relevant observations. The research team will also record attendance at training sessions. These records will be used to assess adherence to the intervention protocol and identify any deviations. VHVs and participating household members will receive modest financial compensation for their time and participation, in accordance with local standards.

We will compare the co-designed Saan Suk intervention with the current practice. Current practice refers to the usual household visits and routine health information provided by VHVs who have received standard training but have not undergone Saan Suk training. The comparator was selected to reflect real-world conditions and to estimate the incremental effect of the Saan Suk intervention beyond standard community-based health communication. To minimise contamination, control clusters will not be exposed to Saan Suk-specific materials, messages, or training content. Participation in the Saan Suk trial does not restrict access to usual health care, routine VHV services, or other public health programmes normally available in the community.

As this trial involves a non-clinical educational intervention, no medical risks are anticipated. However, criteria are defined for discontinuing or modifying the allocated intervention at the individual, provider, and trial levels. For households and individuals, we will discontinue (stop all remaining study visits for that household) if any of the following occur: (1) participant or household head requests to stop participation at any time, for any reason (no justification required); (2) any safety event during or within 24 hours after a visit that poses immediate risk to the participant, household member, or VHV (e.g., physical aggression, threats, severe dog bite or animal attack, hazardous environment); (3) serious adverse event plausibly related to the visit (e.g., injury while acting on a demonstration); (4) confidentiality concern that cannot be mitigated; and (5) loss of eligibility, such as when a participant moves away or lacks capacity to consent. Discontinuation may also occur following a request from VHVs, community leaders, or local authorities. At the VHV level, the intervention delivery will be discontinued if any of the following occur: (1) the VHV fails competency checks after training, (2) breaches confidentiality, or (3) is implicated in a safety-related complaint. In such cases, households will be reassigned to another trained VHV within the same village where feasible. If reassignment is not possible, the intervention will be modified (e.g., delivered in leaflet-only format by a supervisor) or discontinued, and this will be documented as a delivery limitation rather than a crossover.

At the trial level, cRCT may be discontinued (1) in case of major logistical, environmental, political, and civil unrest barriers preventing continuation of the study procedures, (2) ethical or safety concerns raised by the Ethics Committee or relevant authorities, or (3) external events such as natural disasters or public health emergencies that make study implementation unsafe. Discontinuation may also occur if there is (4) an inability to enrol or retain clusters or participants at a sufficient rate to ensure trial completion, (5) loss of implementing partners or VHV network capacity, or (6) data quality failure compromising interpretability, such as when more than 20 to 30% of primary outcome data are missing.

### Primary and secondary outcomes

The primary outcome is self-reported knowledge, assessed as a score on a 23-item structured test covering animal reservoirs and pathogen transmission, preventive behavioural practices, disease ecology, and applied risk-reduction knowledge. Items are scored as binary variables (0/1), with a total possible score of 100, where higher scores indicate greater knowledge. The knowledge test contains single-choice and multiple-choice questions and hierarchical items, i.e., knowledge questions that are asked only if the respondent answered the previous question correctly. This adds a difficulty structure to the knowledge scale and avoids a situation in which respondents with less knowledge have fewer opportunities to get items wrong.

Secondary outcomes (self-reported) include:

- Behaviour, assessed using a 9-item frequency-based scale across wildlife contact, food safety, and hygiene domains. Responses are scored on a 5-point Likert scale, with reverse coding for risk behaviours, and summed to generate a composite score (maximum 36), with higher scores indicating more protective behaviour.
- Attitudes, assessed using 20 Likert-scale items based on Health Belief Model constructs (susceptibility, severity, benefits, barriers, cues to action). Items are scored from 1 (strongly disagree) to 5 (strongly agree), with reverse coding for risk-promoting attitudes, yielding a total score of 100.
- Self-efficacy, assessed using a 7-item scale adapted from the General Self-Efficacy Scale and contextualised to zoonotic risk behaviours. Items are scored on a 4-point Likert scale (1–4), with higher scores indicating greater perceived ability to perform protective behaviours (maximum 21).
- Contacts (presence/absence) with zoonotic hosts are assessed as a contact matrix covering 27 animal species and multiple exposure types (e.g., proximity, handling, environmental contact), recorded as binary variables.

Socio-demographic and experiential variables are collected to describe the study population and adjust for potential confounding. The questionnaire was reviewed by the experts from Thailand and Europe and piloted with a small sample (n=24) of the target population in November 2025, in Khon Kaen Province, Thailand, to assess its clarity and feasibility. Minor revisions were made before field implementation. The questionnaire was developed based on several key publications identified through a literature review, including PREDICT project questionnaires [35,36], WHO-UNICEF WASH household survey [37], and studies on health literacy among pet owners and livestock farmers, mosquito-borne diseases, wildlife trade, and bushmeat consumption in Africa [38] and Southeast Asia [39,40][41,42]. It is divided into five sections: (1) general information and participant ID generation; (2) knowledge, attitudes, self-efficacy, and behaviour related to zoonotic disease and spillover risk; (3) exposure to animal and environmental risks across different sites usually visit (home, temple, garbage dump, water body, agricultural field, and forest) and animal contacts reported; (4) socio-demographic and livelihood characteristics; and (5) contamination and process monitoring questions.

Primary and secondary outcomes will be assessed among recruited household members in both arms during the week following completion of intervention delivery. The Village Heads will invite participants to attend the assessment interview session in the community centre. The trained trial team will obtain informed consent when it has not been obtained previously. The interview will be conducted using a structured questionnaire and will last 45-60 minutes.

### Data management plan

Outcome assessment data will be collected electronically in the field using the Lime Survey platform and securely stored on password-protected servers hosted by Heidelberg University, Germany. Outcome assessors will receive standardised training covering interview techniques, use of electronic data collection tools, ethical conduct, and neutral communication procedures. Training will emphasise the use of standardised wording to avoid any reference to intervention, thereby measuring potential contamination between the arms. Built-in validation checks (e.g., range checks, skip logic, mandatory response fields, and outlier and unusual values) will be embedded in the electronic data collection system. Where internet connectivity is limited, paper-based forms will be used temporarily and subsequently entered into the electronic system following secure data transfer procedures.

Unique identification codes will be assigned to participants, VHVs, and villages. A linkage (key sequence) file connecting identifiers to personal information will be stored separately in a secure, access-restricted location and maintained by the data manager. Data quality will be ensured through built-in validation checks within the electronic data capture system, daily monitoring of incoming data, and periodic review of completed records by the data manager and the field team. The data manager will oversee data cleaning, coding, and verification across all datasets, including quantitative data during the ongoing outcome assessment. Data management and confidentiality procedures will follow institutional and General Data Protection Regulation (GDPR)-compliant standards for the collection, storage, processing, and archiving of personal and research data.

All personal information collected during the trial will be handled in accordance with strict confidentiality procedures before, during, and after the study. Participant data will be pseudonymised at the point of collection using unique study identification codes, and identifiable information (e.g., name and contact information) will be stored separately from research data in secure, access-restricted systems. Only authorised members of the trial coordination team will have access to the linkage file.

All electronic data will be stored on secure, password-protected servers hosted by Heidelberg University. Access will be restricted based on user roles. Paper-based materials, including consent forms, will be stored in locked cabinets at Khon Kaen University, Thailand. All analyses, reports, and publications will use aggregated or anonymised data. No individual participants will be identifiable in any dissemination outputs, including publications, presentations, or any other dissemination materials. All study data will be retained for up to 10 years in accordance with institutional archiving policies and then securely destroyed.

The coordinating centre for the Saan Suk trial is based at the University Hospital Heidelberg (UKHD), Germany, with operational support from Khon Kaen University (KKU), Thailand (Local trial team). UKHD is responsible for overall trial coordination, including regulatory management, ethics approvals, data systems, statistical analysis, and quality monitoring. At the same time, KKU leads day-to-day field implementation and documentation, including coordination with Village Heads and VHVs, scheduling of field activities, training, and engagement with local authorities. A central trial management team comprising researchers and field officers from UKHD and KKU will meet regularly to review progress and coordinate activities.

This trial does not involve a formal data monitoring committee, as Saan Suk is a minimal-risk, behavioural intervention with no clinical dosing, radiation, or invasive procedures. Therefore, no formal interim effectiveness analyses are planned, and anticipated harms are primarily social or operational. Consistent with risk-proportionate oversight, an independent Trial Steering Group (TSG) will provide external monitoring of safety, data quality, contamination, and trial conduct. TSG members who are not involved in intervention delivery or data collection will receive masked, de-identified reports (clusters labelled A/B) and may request unmasking only in exceptional circumstances (e.g., for safety reasons), which will be documented in the minutes.

### Safety considerations

This is a minimal-risk, education-based intervention. Physical medical risks are minimal, the primary foreseeable social or psychological risks (e.g., conflict, stigma, or distress) are minimal; and safety concerns during or after visits (e.g., dog bites while approaching homes) and travelling to the meetings. The intervention trial arm will be monitored through developed adverse event logs.

### Statistical analyses

The statistical analysis will follow the intention-to-treat principle. All eligible and consenting household members selected for outcome assessment will be analysed according to the randomised allocation of their village, regardless of intervention exposure, adherence, protocol deviations, or withdrawal from intervention activities. The village is the unit of randomisation, while outcomes are measured at the individual participant level. All analyses will therefore account for clustering at the village level. The allocation groups will remain masked to the statistician where feasible, until the primary analysis code and outputs are finalised. The primary comparison will be between villages allocated to the Saan Suk intervention and villages allocated to the current practice. All effect estimates will be presented with 95% confidence intervals. Statistical significance will be assessed using two-sided tests with an alpha level of 0.05

A per-protocol analysis will be conducted as a sensitivity analysis. Here, we will include participants in intervention villages who received at least three or four planned visits, and participants in control villages with no documented exposure to Saan Suk-specific materials or training content.

Baseline characteristics will be summarised by cRCT arm to assess the comparability of the randomised groups and to describe the study population. Because randomisation is conducted at the village level, baseline balance will be assessed at both the cluster and individual levels. Cluster-level baseline characteristics include subdistrict, distance to neighbouring study villages, number of registered VHVs, and relevant ecological characteristics, i.e., land use classes (e.g., vegetation, agricultural areas) and land-use interfaces where contact between people and wildlife may occur. Individual-level baseline characteristics will include age, sex, education, income, occupation category, household size, prior experience of zoonotic infectious diseases, and prior awareness of zoonotic diseases. The baseline assessment of the outcomes will not be conducted due to restricted operational feasibility. The background characteristics will be presented as means and standard deviations or medians and interquartile ranges for continuous variables, and as counts and percentages for categorical variables. Cluster size variation will be described and considered in sensitivity analyses.

The primary outcome is the total knowledge score at post-intervention assessment, treated as a continuous variable. Between-arm differences will be estimated using a linear mixed-effects model with identity link, consistent with the assumptions used for the sample size calculation. The model will include a fixed effect for trial arm (pre-training vs post-training) and random intercepts for cluster (village) to account for within-cluster correlation. Pre-specified adjustment covariates include district, VHV ID, age group, sex, education category, and occupational category. Additional adjustment for further baseline variables will be considered if important imbalances are observed despite randomisation.

In addition to the total score, planned stratified analyses of domain-specific knowledge scores will be conducted using the same mixed-effects modelling framework. These analyses are considered exploratory and will not replace inference based on the total knowledge score. The intraclass correlation coefficient will be estimated for the primary outcome.

Secondary outcomes (behaviour, attitudes, self-efficacy, and contacts with zoonotic hosts) will be analysed using mixed-effects regression models appropriate to the scale and distribution of each outcome (e.g., linear mixed models for approximately continuous outcomes; ordinal or generalised linear mixed models for ordinal or non-normal outcomes). For all secondary analyses, cluster will be included as a random effect, and district and VHV will be included as fixed effects where relevant. Effect estimates will be reported with 95% CIs and two-sided p-values. All secondary analyses are exploratory and will be interpreted descriptively. No adjustment for multiple comparisons will be applied.

We do not anticipate substantial missing data due to the following established data collection and quality-control procedures: the data collection tool prompts for answers to each question before proceeding, all interviewers will be trained to complete all items, and data entries will be monitored daily by the field trial supervisor and data manager. If critical items are missing (especially for primary outcomes), participants will be re-contacted within a day. Nevertheless, the extent, patterns, and potential mechanisms of missing data will be systematically examined. If missingness is judged to be missing at random (MAR), missing data will be addressed using multiple imputation. The primary analysis will then be conducted on the imputed or weighted datasets as part of the sensitivity analyses. If missingness is not at random (MNAR), we will conduct sensitivity analyses using MNAR-suitable models, such as pattern-mixture models and delta-adjusted multiple imputation/tipping-point analyses.

Overall, the extent and pattern of missing data will be summarised by trial arm, village, and outcome. Reasons for missing data will be documented where available, including refusal, absence, withdrawal, incomplete questionnaire, loss to follow-up, or data collection error. We will compare participants with complete and incomplete outcome data using baseline socio-demographic characteristics, outcome values where available, village, intervention dose (number of received visits), and trial arm. To assess potential selection bias, we will describe and compare the household selection, consent, refusal, non-response, and loss-to-follow-up rates between trial arms and across clusters. Where information is available for non-participants or participants lost to follow-up, we will compare them with included participants to assess whether missingness or non-participation may be associated with allocation group or key baseline characteristics.

### The status and timeline of the study

The trial is expected to start on the 27th of July 2026 and continue until October 2026. Below, we present the participant timetable (Table 2). This article reports the initial protocol approved by the Ethics Committee of Heidelberg University Hospital (S-607/2025, 14.04.2026), before recruitment. Recruitment of villages, VHVs, and households is planned to start on 27 July 2026 and be completed on 16 August 2026. The intervention implementation will start on 17 August 2026 and last for four weeks. Outcome assessment will be conducted between 14 and 30 September 2026 in both trial arms. Database lock is planned for November 2026. No participants have been enrolled at the time of submission. A manuscript reporting the main trial results is expected to be submitted for peer review in the third quarter of 2027, when the trial results will become available.

**Table 2:**
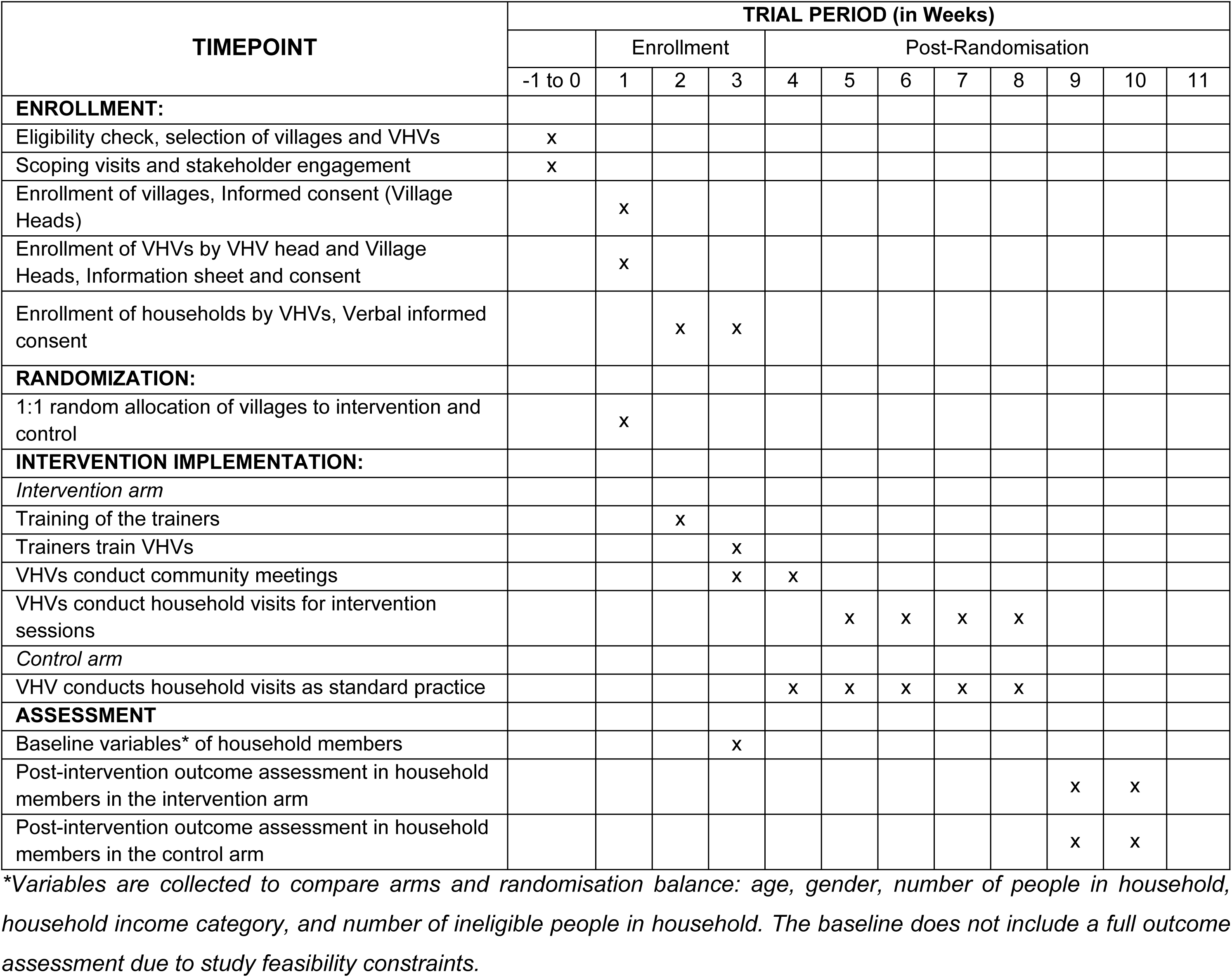
Schedule of enrollment, interventions, and assessments for the Saan Suk Trial.

### Data availability

The pseudonymised participant-level dataset will not be made publicly available. De-identified individual participant data (IPD) underlying the primary and secondary outcomes can be shared after study completion and publication of the main results. Data will be available to qualified researchers upon request and approval of a data use agreement. Shared materials will include the final dataset, data dictionary, and study protocol. No identifiable information will be released, and all data sharing will comply with ethical and data protection regulations. Data use is therefore restricted to analyses outlined in the Saan Suk trial protocol and approved by the Ethics Committee. Statistical code and data dictionaries will be retained within the trial coordination team and may be made available upon reasonable request, subject to approval by the principal investigator and in accordance with ethical, GDPR, and institutional data protection requirements.

## Discussion

In this cluster randomised controlled trial, we will evaluate the effects of a co-designed One Health educational intervention delivered through Thailand’s existing Village Health Volunteer system in rural communities in Chanthaburi Province. The co-design approach is intended to ensure that the intervention is desirable, acceptable, and feasible within the target communities of Thailand. By leveraging existing VHV training and community health structures supported by the Ministry of Public Health, the Saan Suk programme has the potential to be integrated into the routine VHV activities if it is shown to be effective and feasible.

The choice of cluster randomisation is grounded in the nature of the intervention and its intended mode of delivery. If scaled up, the Saan Suk intervention would be implemented at the village level through registered VHVs, rather than delivered to isolated individuals. Village-level randomisation, therefore, reflects the expected implementation pathway and supports community acceptance and feasibility of the randomisation procedures. It also reduces the risk of contamination between participants within the same community. Nevertheless, intervention spillover (contamination) between trial arms remains possible because of participant mobility and close communication with family members, friends, and neighbouring communities. To reduce this risk, villages must be located at least 2 km apart to be eligible for inclusion in this trial, and community engagement meetings will emphasise the importance of not sharing study-specific procedures, materials, or content with neighbouring villages during the study period. The distance for the restriction was chosen to be feasible for selecting the required number of villages and based on expert opinion on residents’ travel frequency. Potential contamination will also be assessed through selected questions in the outcome assessment questionnaire.

We acknowledge that the implementation of the Saan Suk programme depends on the availability, motivation, and capacity of VHVs. VHVs are trusted community actors well placed to deliver household-level health education interventions; however, they may differ in experience, communication style, time availability, and confidence in delivering One Health content. To support intervention fidelity, VHVs in the intervention arm will receive standardised training during a two-day workshop, including a manual, structured materials, and communication training. Adherence and fidelity will be monitored through visit documentation, conducted by VHVs and evaluated by the trial coordination team.

Village Heads will also play an important role in supporting trial implementation, including encouraging adherence to training and field procedures. Their involvement in the allocation process, including the public envelope draw, is intended to strengthen transparency, community acceptance, and local ownership. Village Head permission is necessary for the respective village participation in the study; however, individual informed consent will also be obtained from trial participants, including VHVs and adult household members, where they participate in study-specific procedures or outcome assessment.

Outcome assessment is based on self-reported information, including knowledge, attitudes, self-efficacy, behaviours, and contacts with local animal hosts. These outcomes are appropriate for evaluating behavioural determinants of zoonotic spillover prevention, but they are subject to self-report and social desirability bias, particularly after delivery of an educational intervention [43]. To reduce measurement bias, outcome assessors will be trained, standardised questionnaires will be used, and outcome assessment teams will remain blinded to the intervention allocation. The questionnaire was expert-reviewed, back-translated, and piloted in the target communities to improve clarity, cultural appropriateness, and acceptability. The questionnaire also includes items to collect information on potential baseline factors that may differ between the trial arms and, if necessary, can be adjusted for in the analysis. Non-self-reported outcomes of zoonotic disease prevention behaviour or spillover risk are not feasible within the scope of this trial.

Participant retention and complete follow-up may be affected by livelihood patterns, agricultural work, seasonal mobility, weather, and competing community activities. Activity logs will be conducted regularly by the trial coordination team. Field teams will document reasons for non-response, discontinuation, or missing data, and the analysis will account for clustering and missingness in accordance with the pre-specified statistical analysis plan.

Finally, the trial will be conducted in a specific ecological, cultural, and health-system context. The findings will be most directly applicable to rural Thai communities where VHVs are active and where community members regularly interact with wildlife, domestic animals, and shared environments. However, the intervention development process, delivery model, and evaluation framework may be relevant to other settings aiming to strengthen community-based pandemic prevention and One Health literacy through existing community health structures. Results will be disseminated through peer-reviewed open-access publications and presentations at scientific conferences, irrespective of study outcome. Findings will also be shared with local and national stakeholders in Thailand, including community feedback sessions where appropriate. A summary of results will be made available to study participants and communities in an accessible format (e.g., via the project website). Trial results will be reported in the relevant trial registry to the responsible ethics committees, in accordance with applicable reporting requirements.

Any protocol modifications or amendments will be submitted to the relevant ethics committees for approval before implementation. Approved amendments will be communicated to the trial registry, investigators, and study staff. All deviations from the protocol during trial implementation will be documented in a protocol deviation log, reviewed by the trial coordination team, and considered in the analysis where relevant.

Overall, the Saan Suk trial addresses an important evidence gap regarding whether a co-designed, theory-informed, VHV-delivered One Health intervention can improve community knowledge, behavioural determinants, and self-reported practices related to zoonotic disease prevention, thereby contributing to the broader pandemic prevention agenda.

## Abbreviations

CI: Confidence interval
cRCT: Cluster Randomised Controlled Trial
EIDs: Emerging Infectious Diseases
FGD: Focus Group Discussion
GDPR: General Data Protection Regulation
HCD: Human Centred Design
HBM: Health Belief Model
ICC: Intracluster Correlation Coefficient
ID: Identification Number
ITT: Intention-to-treat
MAR: Missing at Random
MNAR: Missing Not At Random
MOPH: Ministry of Public Health
PAO: Provincial Administrative Organisation
PI: Principal Investigator
SD: Standard deviation
THB: Thai Baht (currency)
TSG: Trial Steering Group
VHV: Village Health Volunteer

## Declarations

## Acknowledgements

We thank Three O’Clock for contributing to the design elements of the Saan Suk intervention and Mariya Makarova for video production. We also gratefully acknowledge Dr. Montree Prasertrungruang and the staff of the KNOWA Learning Complex in Kranuan District, Khon Kaen Province, for their support in organising the pilot of the intervention.

## Authors’ contributions

Conceptualisation: MT, TB, JR, HJO;

Study design: MT, CPR, SC, PP, MP, TB, JR, HJO, PL;

Intervention co-development: MT, CRP, PP, SC, KB, UK, MP, HJO, PL;

Community engagement and local implementation planning: MT, CRP, PP, SC, KB, UK, MP, HJO, PL;

Trial coordination: MT, CRP, SC, MP, HJO, PL;

VHV training materials and field procedures: MT, CRP, UK, PP, SC, KB, MP, HJO, PL;

Sample size calculation: MT, CRP;

Statistical analysis plan: MT, JR, TB, HJO;

Data management procedures: MT, CRP, SC;

Ethics and regulatory documentation: MT, CRP, UK, JR, TB, KJ; PL;

Writing – original draft: MT, CRP;

Writing – review and editing: All authors

Funding acquisition: MT, TB, JR, HJO

All authors reviewed and approved the final manuscript and agreed to be accountable for the work.

## Ethics approval

This study has been approved by the Ethics Commission of the Medical Faculty of Heidelberg University (reference number: S-607/2025, version 3 from 31 March 2026) and is registered in the DRKS registry (Registration ID: <u>DRKS00038582</u>; date of registration: 11 May 2026). Before enrollment, all participants are informed about the study’s objectives and their right to withdraw at any time without any negative consequences, and provide written informed consent.

## Appendix

S1 Table. SPIRIT 2025 checklist of items to address in a randomized trial protocol

